# Sword of Damocles or choosing well. Population genetics sheds light into the future of the COVID-19 pandemic and SARS-CoV-2 new mutant strains

**DOI:** 10.1101/2021.01.16.21249924

**Authors:** J. G. García de Alcañíz, V. López-Rodas, E. Costas

## Abstract

An immense scientific effort has been made worldwide due to Covid-19’s pandemic magnitude. It has made possible to identify almost 300,000 SARS-CoV-2 different genetic variants, connecting them with clinical and epidemiological findings. Among this immense data collection, that constitutes the biggest evolutionary experiment in history, is buried the answer to what will happen in the future. Will new strains, more contagious than the current ones or resistant to the vaccines, arise by mutation? Although theoretic population genetics is, by far, the most powerful tool we have to do an accurate prediction, it has been barely used for the study of SARS-CoV-2 due to its conceptual difficulty. Having in mind that the size of the SARS-CoV-2 population is astronomical we can apply a discrete treatment, based on the branching process method, Fokker-Plank equations and Kolmogoroff’s forward equations, to calculate the survival likelihood through time, to elucidate the likelihood to become dominant genotypes and how long will this take, for new SARS-CoV-2 mutants depending on their selective advantage. Results show that most of the new mutants that will arise in the SARS-CoV-2 meta-population will stay at very low frequencies. However, some few new mutants, significantly more infectious than current ones, will still emerge and become dominant in the population favoured by a great selective advantage. Far from showing a “mutational meltdown”, SARS-CoV-2 meta-population will increase its fitness becoming more infective. There is a probability, small but finite, that new mutants arise resistant to some vaccines. High infected numbers and slow vaccination programs will significantly increase this likelihood.

## Introduction

Due to Covid-19’s pandemic magnitude, an immense effort to sequence SARS-CoV-2 genetic variants and its relations to clinical and epidemiological findings has been made worldwide. There are nearly a thousand preprints available about genetic variants, published in https://www.medrxiv.org ^1^, https://www.biorxiv.org^2^ and some other preprint platforms, which promote the rapid data exchange about SARS-CoV-2 genetic sequences^3–10^, infectivity and lethality (https://nextstrain.org/sars-cov-2/; https://www.ncbi.nlm.nih.gov; https://www.gisaid.org; https://covid.pages.uni.lu; https://www.covid19dataportal.org)^11–15^. Updated mutations on any sequence can be tested in different countries and time periods.

There is an impressive descriptive knowledge of all these mutants at a molecular level. The relation between the genetic sequence, the protein coded and its role in the infectivity mechanism, so its potential infectivity can be inferred some examples of this are explained in Wang et al., 2020; Islam et al., 2020; Brufsky, 2020; or Bajaj & Purohit, 2020^16–19^.

Some surprising enigmas around SARS-CoV-2’s population genetics are revealed thanks to these enormous experimental efforts. First of these enigmas appears when comparing current genomes in world’s population with the original strain isolated in Wuhan. Almost 300,000 genetic variants of the SARS-CoV-2 have already been detected since then, such variability is shocking.

It is not the only one. The origin of new strains like for example B117 (also known as VUI202012/01) is also extremely surprising. This new strain accumulates 23 different mutations, of which 17 happened abruptly at the same time. Many are spike protein mutations (i.e., 69-70, deletion 145, N501Y, A570D, D614G, P681H, T716I, S982A, D1118H) that make it much more infective than the ancestral strain. This mutant variant become dominant in Southern England, it is striking the UK and is starting to spread around the globe.

The likelihood of suddenly having 17 new mutations in a new strain is incredibly low. The likelihood that without recombination, even with mutation rates much higher than the mutation rate observed in SARS-CoV-2, seventeen new mutations accumulate in a virus tends to zero. A simple calculation assuming a 10^−3^ mutation rate (significantly higher than SARS-CoV-2 mutation rate) shows the unlikeliness of this event.

Furthermore, the likelihood of all these mutations resulting in a more transmissible strain, again, is close to zero. It should never had happened. However, according to a classical viral evolution perspective, with time new mutant strains should be less and less effective due to Muller’s rachet and mutational meltdown^20–22^.

Mutation is a random event and most of these mutations are deleterious or neutral. In an asexual population (in absence of recombination) these deleterious mutations with time accumulate, and the population may become extinct, precisely because the accumulation of these deleterious mutations. Muller’s rachet has been demonstrated, at a theoretical and at an experimental level in many RNA viruses, and many other organisms without recombination^23–27^.

With this setting it does not come as a surprise that it was thought that with time, SARS-CoV-2 meta-population would suffer mutational meltdown. It was even suggested to increase SARS-CoV-2 mutation rates to fight the virus^28–30^.

Then, how is it possible that a strain that accumulates 23 mutations, of which 17 are new, has much more biological effectiveness than the strain it originates from? and, why is it possible that as time goes by, SARS-CoV-2 meta-population is more effective?

Is a fact that new mutant SARS-CoV-2 strains detected spreading among populations have a significantly higher biological effectiveness than the ancestral strains from which they originate. A quick look to databases shows that new mutant strains continue to emerge and that they are more infective than the ancestral strains.

The time has come for the theoretical population genetics to give an answer. We must be aware that, from a scientific perspective, we are at the biggest evolution experiment in history. A virus, that has colonized the enormous ecological niche of human race and has expanded to such level that its numbers are astronomical. In population’s terms is a population consisting of an infinite but “countable” number of individuals^31^. It never happened before, such an interesting evolution phenomenon that can be scientifically studied as it happens. With the amount of data gathered we are before an extraordinary opportunity to analyse live SARS-CoV-2 evolution^10,32–34^.

Furthermore, from a practical point of view, population genetics can do accurate predictions about where SARS-CoV-2 evolution goes and what the consequences may be.

We will indicate that observing such variability within SARS-CoV-2 meta-population nor the arising of even more infective strains (as B117 or 501.V2) is something rare. Theoretical population genetics predicts it.

Most likely it will happen again, a new strain could emerge in the near future with even more infectious ability than B117 or 501.V2.

### Fixation and extinction dynamics of new mutants in the SARS-CoV-2 meta-population

We know, since the seminal paper of Luria & Delbruck (1943)^35^ with *α*-bacteriophage and its host *E. coli*, that the emergence of mutants is a recurrent process that happens randomly, pre-selectively and pre-adaptatively. There will constantly be arising all different new mutants in the SARS-CoV-2 meta-population, even the same mutation will happen now and again. The key to what is happening with new mutations that originate new SARS-CoV-2 strains is to figure out the fate of this new mutants. For that, three issues have to be tackled:

1. Calculate the probability for a new SARS-CoV-2 mutant to survive through time depending on its selective advantage or disadvantage.
2. Calculate the probability for a new SARS-CoV-2 mutant to fixate or become the dominant genotype in a SARS-CoV-2 population depending on its selective advantage or disadvantage.
3. Calculate time needed for a newly arisen mutant to increase its frequency to become dominant in the SARS-CoV-2 population depending on its selective advantage or disadvantage.

We will mathematically solve these uncertainties and draw the consequences.

#### 1. Probability that a new SARS-CoV-2 mutant newly arisen in the SARS-CoV-2 population survives through time depending on its selective advantage or disadvantage

We will aboard this problem with a discrete approach, based on the branching-process method in a population of infinite but “countable” individuals developed by Crow & Kimura (1970)^31^ based on Haldane (1927)^36^ and Fisher’s (1930a, 1930b)^37^ first works.

Being a new mutant virus newly arisen in the SARS-CoV-2 population, let *p*_0_, *p*_1_, *p*_2_, …*p*_*k*_ be the probabilities that the new mutant virus will become 0, 1, 2, … *k* in number in the next generation.

*p*_0_ is the probability that the new mutant will be lost in the next generation (0 < *p*_0_ < 1). The probability generating function *f*(*x*) is:

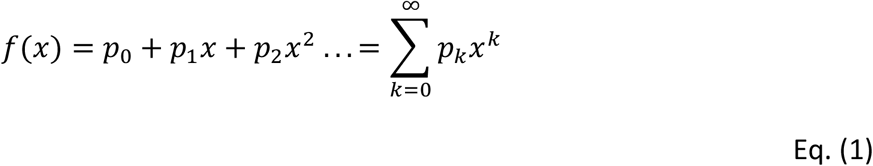

In the next generation the mutant viruses reproduce independently so that each mutant virus again leaves *p*_1_, *p*_2_, … *p*_*k*_ probabilities that will become 0, 1, 2, … *k* in the next generation. So: 

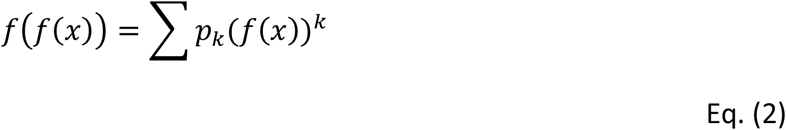

This reproduction pattern continues, so the probability-generating function for the number of mutant viruses after *t* generations is given by the *t* interation of the *f*(*x*): *f*(*f*(*f* … *f*(*x*))) which can be expressed by *f*_*t*_(*x*). So: 

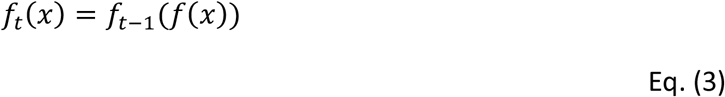

The probability that the virus is lost by the *t* generation (*v*_*t*_) is given by *f*_*t*_(0) 

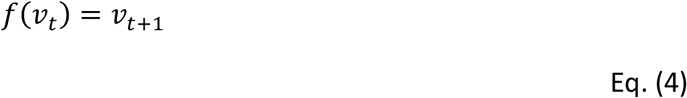

Now we must assume a probabilistic distribution for the number of descendants of a virus, Poisson distribution is both easiest and realistic, in this way: 

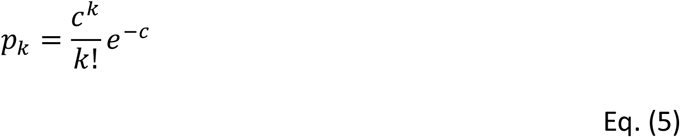

were *c* is the average number of descendant mutant viruses left in the next generation. Then the probability generating function -Eq. (1)- for this distribution is: 

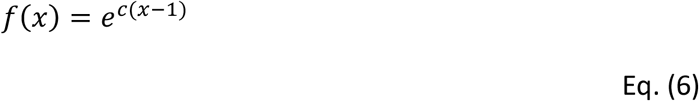

In the event that natural selection acts on the new mutant virus, then *s* is the selective advantage of the mutant virus. If new mutation is disadvantageous, neutral or advantageous then *s* < 0, *s* = 0, or *s* > 0 respectively.

Approximately, *s* = *c* − 1 and for advantageous mutant viruses (*c* > 1) the probability of survival *u* is: 

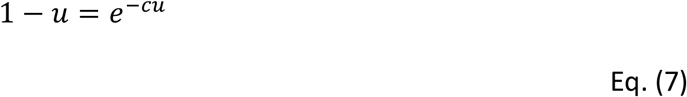

Based on equations of this branching-process method in a population of infinite but “countable” viruses, we estimate the survival likelihood for mutant strains with a 10% selective disadvantage (*c* = 0.90), a 5% selective disadvantage (*c* = 0.95), neutral mutant strains (*c* = 1.00), and for advantageous mutant strains with 5% more advantage (*c* = 1.05), 10% more (*c* = 1.10), and a 50% more (*c* = 1.50) (Figure 1).

**Figure 1.**
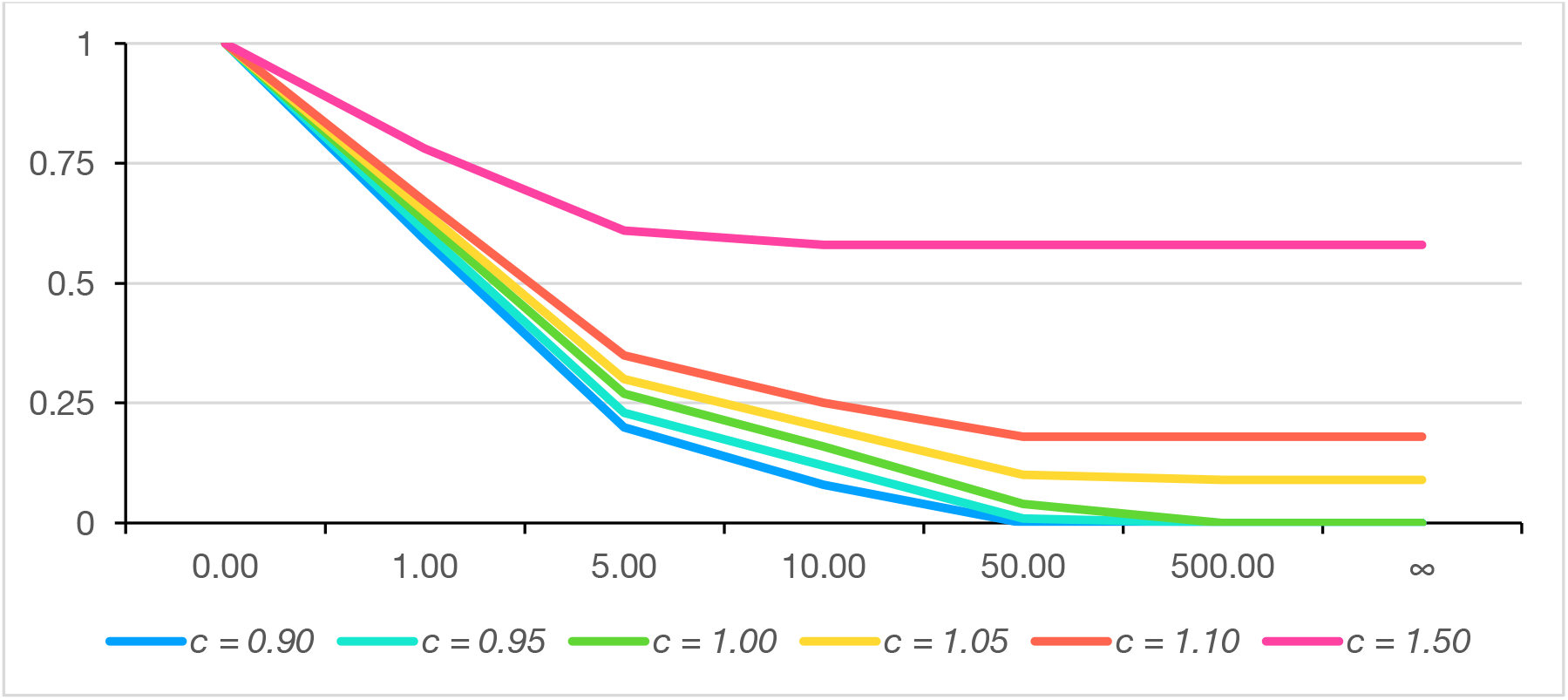
Survival likelihood for neutral, deleterious and advantageous SARS-CoV-2 mutants estimated from u_t_ = 1 – e^-cu^and u_0_ = 1.00

It is clear that all mutants with a selective disadvantage will never fixate and will be lost soon enough, usually before the 50^th^ viral generation.

Neutral new mutants (i.e., with no selective advantage or disadvantage) neither will survive long in the SARS-CoV-2 population. A neutral mutant has only a 63% chance of surviving the next generation and only of a 16% after 10 generations. After 50 generations, a new neutral mutant has only a 4% chance of remaining in the population. In one hundred generations the chances drop below 2%. Taking in consideration the viral growth kinetics this happens in a very short period of time.

Even mutants with very small selective advantages will have very low survival likelihoods. With a 5% selective advantage (which is quite considerable) the survival likelihood only climbs to a 10% after 50 generations. A 10% advantage (really high) only gives an 18% survival likelihood in the 50^th^ generation. Small selective advantages, usually happening in the best mutants, barely change the survival likelihood.

Enormous selective advantages are needed for the survival likelihood to increase significantly after some generations. In this sense, a new mutant with a 50% selective advantage (something remarkably extraordinary) has a 78% survival likelihood after one generation, a 60% in the second and of 58% in the 50^th^ generation.

However, all mutants with a selective advantage show an interesting phenomenon. The survival likelihood keeps falling generation after generation up to a point at which it remains constant (Figure 1).

#### 2. Fixation likelihood of a newly arisen mutant, within the SARS-CoV-2 population, depending on its selective advantage or disadvantage

Wright (1945)^38,39^, Kimura (1957, 1962)^40,41^ and Crow & Kimura (1970)^31^ introduced the use of Fokker-Plank equations in population genetics theory. Here we use this procedure employing the Kolmogoroff’s forward equations^42^ to calculate the fixation likelihood of a new mutant SARS-CoV-2 strain depending on its selective advantage.

We assume that the size of the SARS-CoV-2 population is big enough as to consider the change frequency process of the mutant strain through time as a continuous stochastic process.

The likelihood *u*, of a new SARS-CoV-2 mutant strain to be fixed within the population in the *t*^th^ generation if the initial frequency is p, in the t_0_ generation is: 

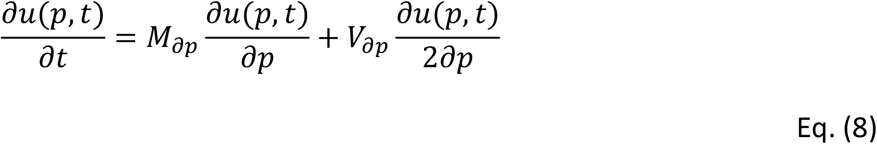

were *M*_*dp*_ and *V*_*dp*_ are the median and the variance of the frequency change *p* for the mutant strain per generation.

The likelihood *u*(*p, t*) is calculated using partial differential equations with boundary conditions: 

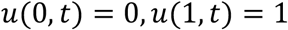

The ultimate probability of fixation is defined by: 

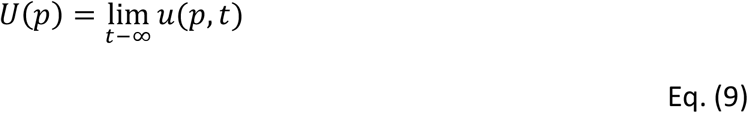

and by: 

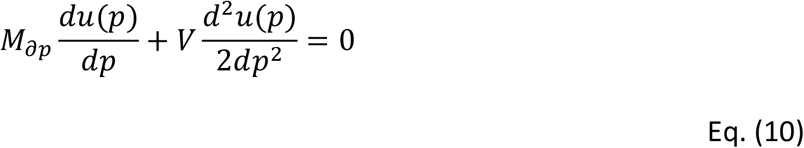

Assuming that the progeny number of new mutant strain follows a Poisson distribution 

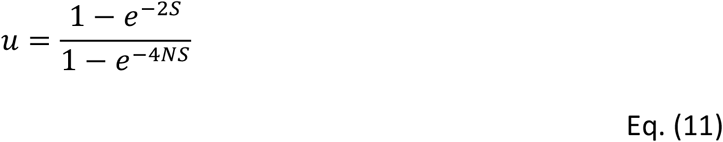

Employing the Kolmogoroff’s forward equations in a population of infinite but “countable” virus, we estimate the fixation likelihood within the SARS-CoV-2 population for mutant strains with a 10% selective disadvantage (*c* = 0.90), a 5% selective disadvantage (*c* = 0.95), neutral mutant strains (*c* = 1.00), and for selective advantageous mutant strains with 5% more advantage (*c* = 1.05), 10% more (*c* = 1.10), and 50% more (*c* = 1.50) (Figure 2).

**Figure 2.**
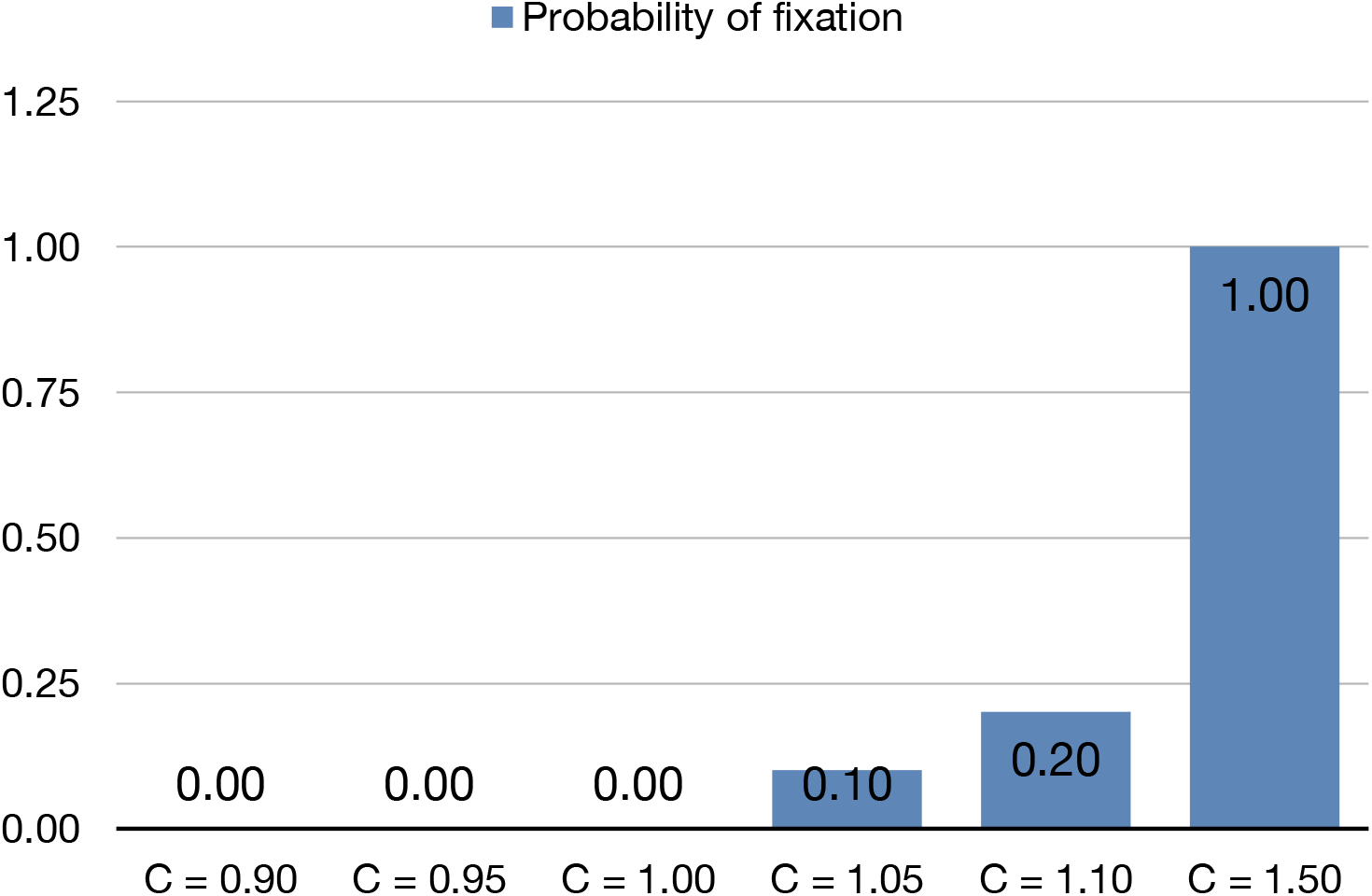
Likelihood for a new mutant to fix in the SARS-CoV-2 population for diferent selective advantages or disadvantages

It is clear that all mutants with a selective disadvantage will never fixate within the SARS-CoV-2 population, neither will neutral new mutants (i.e., with no selective advantage or disadvantage).

Even mutants with small selective advantages will have very low fixation likelihoods. With a 5% selective advantage (which is quite considerable) the fixation likelihood only climbs to a 10%. A 10% advantage (really high) only gives an 18% fixation likelihood. Small selective advantages, usually happening in the best mutants, barely change the fixation likelihood in the SARS-CoV-2 population.

For a newly arisen mutant to fixate within the SARS-CoV-2 population enormous selective advantages are needed (Figure 2).

#### 3. Time needed for a newly arisen mutant to increase its frequency to become dominant in the SARS-CoV-2 population depending on its selective advantage or disadvantage

We assume that in the human population each individual is only infected with one SARS-CoV-2 strain (genotype), that replicates and then spreads out to infect new individuals.

Selection occurs when one genotype leaves a different number of progeny than another. In practice this will be greatly related to the ability each genotype has to infect new individuals. In such way, the key is to estimate the change in frequency of the different genotypes on each infective step (i.e., from one host to the next ones). To simplify notation each infective step within humans we will call it “generation” (Figure 3).

We assume different genotypes *A*_1_, *A*_2_, *A*_3_, … within the SARS-CoV-2 population, which have fitness *w*_1_, *w*_2_, *w*_3_, … and are present in the SARS-CoV-2 meta-population with frequencies *q*_1_, *q*_2_, *q*_3_, …

Let’s suppose that *A*_1_ genotype is the new mutant; from which we want to calculate the gene frequency change. The proportion of A_1_ in the next generation will be: 

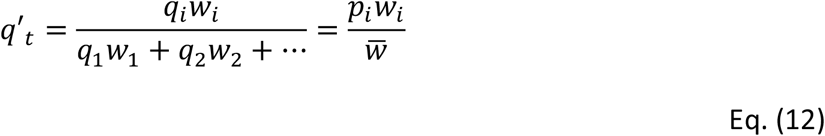

where 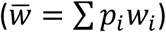 is the average fitness.

Change in *A*_*i*_’s proportion after one generation is: 

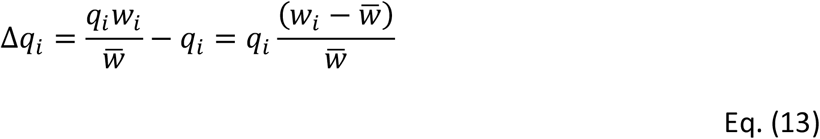

The quantity 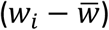 is the average excess in fitness of the genotype *A*_1_.

Consider now time in infective leaps within humans, assuming a continuous model. The change rate can then be visualized as a continuous function 

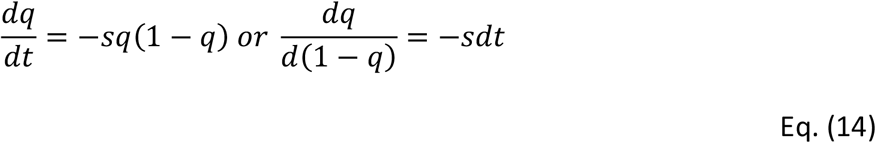

integrating both ends 

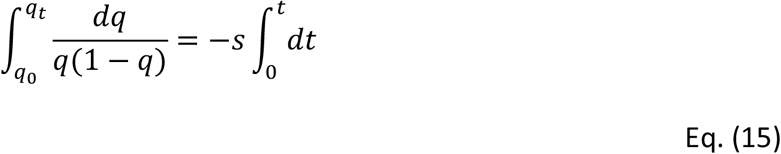

considering *t* as the number of infective steps (i.e., generations) for the frequency of *A*_1_ mutant to change from *q*_0_ to *q*_*t*_, then: 

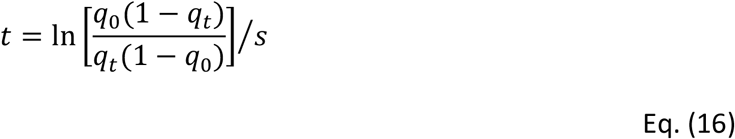

where *s* = *c* − 1 and for advantageous mutant viruses (*c* > 1), number of infective steps (i.e., generations) for the frequency of *A*_1_ mutant to change from *q*_0_ to *q*_*t*_ is: 

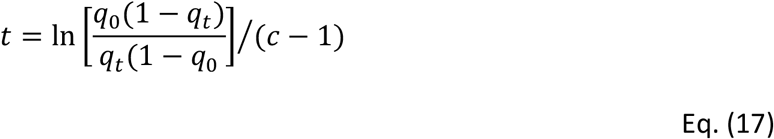

Considering three different settings we estimate the number of transmissions human-to-human will a new mutant SASR-CoV-2 strain, - with a 1% (*c* = 0.01), a 5% (*c* = 1.05), a 10% (*c* = 1.10), a 50% (*c* = 1.50) and a 100% selective advantage (*c* = 2.00) - need to increase its frequency within the SARS-CoV-2 meta-population. The three different settings considered were: i) how many transmissions human-to-human are needed to go from a very low frequency (*q*_0_ = 0.01) to an also a low frequency but easily detectable (*q*_*t*_ = 0.10); ii) How many transmissions are needed to go from a very low frequency (*q*_0_ = 0.01) to be dominant (*q*_*t*_ = 0.51) in the meta-population; iii) how many transmissions needs a new mutant strain to go from a very low frequency (*q*_0_ = 0.01) to be virtually fixed in the meta-population (*q*_*t*_ = 0.99) (Table 1).

**Table 1.** Number of transmissions human-to-human needed for a new mutant to increase its frequency in the population depending on its selective advantage, as well as time needed to achive it on a quick and slow setting

| Number of steps needed to go from $q_0$ to $q_t$: | Selective advantage | | | | |
| --- | --- | --- | --- | --- | --- |
| | $c = 1.01$ | $c = 1.05$ | $c = 1.10$ | $c = 1.50$ | $c = 2.00$ |
| $q_0 = 0.01$ to $q_t = 0.10$ | 240 | 48 | 24 | 5 | 2 |
| $q_0 = 0.01$ to $q_t = 0.51$ | 458 | 92 | 46 | 9 | 5 |
| $q_0 = 0.01$ to $q_t = 0.99$ | 919 | 183 | 91 | 18 | 9 |

There is a lot of uncertainty when estimating the time these transmissions would take. There is great variability not only about the moment a newly infected host starts spreading the virus but also for how long it does. Furthermore, one way a new SARS-CoV-2 mutant could increase its infectivity could be by reducing the time needed for a host to start infecting other people, another could be increasing the host’s infectivity period. In that way, in order to translate number of transmissions into time, we consider a quick setting (Table 2), with 11.5 days as the average time for one human-to-human transmission, and a slow setting (Table 3), with an average 20 days for this to happen. Real time should be between both settings.

**Table 2.** Time needed (in days) for a new mutant to increase its frequency in the population depending on its selective advantage, quickest scenario.

| Time needed (days)<br>Quickest scenario | Selective advantage |  |  |  |  |
| --- | --- | --- | --- | --- | --- |
| | $c = 1.01$ | $c = 1.05$ | $c = 1.10$ | $c = 1.50$ | $c = 2.00$ |
| $q_0 = 0.01$ to $q_t = 0.10$ | 2,760 | 552 | 276 | 57.5 | 23 |
| $q_0 = 0.01$ to $q_t = 0.51$ | 5,267 | 1,058 | 529 | 103.5 | 57.5 |
| $q_0 = 0.01$ to $q_t = 0.99$ | 10,568 | 2,104 | 1,046 | 207 | 103.5 |

**Table 3.** Time needed (in days) for a new mutant to increase its frequency in the population depending on its selective advantage, slowest scenario.

| Time needed (days)<br>Slowest scenario | Selective advantage |  |  |  |  |
| --- | --- | --- | --- | --- | --- |
| | $c = 1.01$ | $c = 1.05$ | $c = 1.10$ | $c = 1.50$ | $c = 2.00$ |
| $q_0 = 0.01$ to $q_t = 0.10$ | 4,800 | 960 | 480 | 100 | 40 |
| $q_0 = 0.01$ to $q_t = 0.51$ | 9,160 | 1,840 | 920 | 180 | 100 |
| $q_0 = 0.01$ to $q_t = 0.99$ | 18,380 | 3,660 | 1,820 | 360 | 180 |

It is important to note that the original SARS-CoV-2 original strain isolated in Wuhan which unleashed COVID-19 pandemic started over a year ago. Since then, we have seen dominant strains, like Wuhan strain during first wave, being replaced by new mutant strains only originated two or three months before their dominance (e. g. strain arisen in Spain during the summer of 2020, or the English B117 and the South African 501Y.V2 emerged around November - December 2020). As seen in Tables 1, 2 and 3, these quick increase in frequencies of some of the new SARS-CoV-2 mutant strains can only be achieved with immense selective advantages (*c* = 1.5 values or higher). There would not have been enough time for these new mutant strains to spread if these tremendous selective advantages were not present.

## Discussion

### 1. The biggest evolution experiment in history

Theodosius Dobzhansky, in 1973, published his paper *“Nothing in Biology makes sense except in the light of evolution”*^43^. This title alone summarises the essence of modern biology thinking, even immersed in the COVID-19 pandemic whirlpool we cannot forget it.

In fact, we are living the greatest evolution experiment in history. Through mutation, a coronavirus has evaded the species barrier and expands uncontrolled in a new colossal ecological niche, the human race.

In over a year nearly 300,000 new mutations of the virus in the SARS-CoV-2 meta-population have been detected. This immense genetic variability, many of which is neutral, changes very quickly in time and space and fluctuates stochastically through genetic drift.

Mutants that increase significantly viral infectivity also emerge. These mutants increase their frequency in the SARS-CoV-2 meta-population favoured by natural selection. The history of successful mutant strains (i.e., S477N, N439K or N501Y) that seem to be more infective and become significantly more frequent in the population, can be traced (https://www.gisaid.org)^13^. D614G strain, for example, emerged at the beginning of the COVID-19 pandemic in Europe, increased its frequency and was responsible of many infections during the first wave. A222V strain, that arise during the 2020 summer spread through Europe and increased its frequency during the second wave. At the end of 2020, B117 strain, defined by multiple spike protein mutations, become dominant in south east England and started spreading around the globe, In November the novel variant 501Y.V2, with triple spike receptor binding substitutions, was detected in South Africa and seems to be also increasing its frequency.

The clue to what is happening to SARS-CoV-2 and can answer some of the important questions we are asking ourselves is in evolution in action. Will much more infective new SARS-CoV-2 variants emerge? Will mutants capable of evading the vaccine arise?

If we want to know what will happen, we must study SARS-CoV-2’s evolution. We have not finished yet with Dobzhansky’s brilliant idea.

### 2. Emergence and fate of new mutations in the SARS-CoV-2 meta-population

Since Luria & Delbruck (1943)^35^, Newcombe (1949)^44^, Novick & Szilard (1950)^45^ and Lederberg & Lederberg’s (1952)^46^ seminal papers, a great number of studies show that mutant emergence is a recurrent process that happens randomly and in a pre-selective and pre-adaptative way (i.e., mutants emerge before natural selection acts)^47,48^. This surprising process has been demonstrated: in microorganism populations, spontaneous and recuring mutant strains arise that have resistance to newly synthetised substances, even before industry developed them^49,50,59,51–58^ even if these substances are of military use and where never before present in nature^60^ or are radioisotopes of nuclear industry^61–64^.

Key in the modern evolution thinking is that evolution forces, as natural selection or genetic drift, act either increasing the frequency of these new mutants within the population or removing them^43,65–67^. Majority of these mutants are deleterious and natural selection rules them out, but since mutation is a recurrent process, they will arise time after time remaining at a very low frequency due to a mutation-selection equilibrium^31,68^. Other mutants are selectively neutral, natural selection does not act upon them and remain balanced through mutation-drift^31,68^. If in the event of a drastic change in the environment, as happens with industrial and mining catastrophic spillages, volcanic eruptions, etc. these strains can be favoured by the new environmental conditions and become dominant in the population^69–75^.

In the SARS-CoV-2 meta-population is easy to anticipate that an enormous number of new mutants, randomly and recurringly, are spontaneously arising (considering that the SARS-CoV-2 mutation rate is higher than 10^−4^, an infected host can produce at least 10^10^ new viruses, in the world there are many millions of infected people). Certainly, among these new mutants some can arise that are more infective than the current ones, even mutants resistant to the already developed vaccines. This is more so in the middle of an infectious wave, when SARS-CoV-2 meta-population grows to a size that even the most improbable mutant can happen. Loss or fixation of these new mutants within the SARS-CoV-2 meta-population will decide the future.

Haldane (1927)^36^, Fisher (1930a, 1930b)^37,76^ and Wright (1931)^77^ were the first ones to significantly contribute to the problem of fixation or loss of new mutants in the populations. In 1957, Kimura^40^, approached this problem using a diffusion model that allowed him to estimate new mutant’s fixation likelihood depending on the population size and selection coefficient. Kimura (1962)^41^ also analysed the probabilities of eventual fixation of a new mutant and the mean and variance of the rate of change of gene frequency per generation. This process was extensively revised by Crow & Kimura (1970)^31^.

Results are clear, most of the new mutants will disappear soon after arising. Only some will succeed increasing their numbers and remaining within the SARS-CoV-2 population for a long time. Failure or success of a new mutant relies at great length on chance and, of course, on natural selection. But natural selection in itself cannot assure the survival of a new mutant even possessing a selective advantage, at least not until it becomes sufficiently abundant in the population^31,78–80^.

Chance can make, in very small populations, that neutral mutants increase their frequency and even fixate in the population, but in such an enormous population as SARS-CoV-2 is, it does not currently happen. As Shown in Figures 1 and 2, none of the new SARS-CoV-2 mutants with a selective disadvantage will never fixate. Neither will neutral new mutants. For a new mutant to increase its frequency in the SARS-CoV-2 population it needs a selective advantage. Even then, mutants with small selective advantages will have very low fixating likelihood. With a 5% selective advantage (quite appreciable) merely have a 10% fixation likelihood. With a 10% selective advantage (really big) fixation likelihood will only go up to 20%. Small selective advantages, usually in some mutants, barely increase their fixation likelihood within the SARS-CoV-2 population.

Only mutants with a really big selective advantage will significantly increase their numbers in the SARS-CoV-2 meta-population. While our progress in medicine and epidemiology makes more effective our fight against SARS-CoV-2, the new mutants arising in the meta-population will be more infective, we reach a Red Queen dynamic^81–83^. We improve our fight against the virus, but it quickly evolves in a continuous adaptation that allows to maintain the status quo.

However, vaccination can radically change this scenario. Antibodies generated by the vaccines target spike proteins and is quite probable that vaccines remain effective against many of these highly infective mutants. Selection changes if vaccination is massive. Highly infective strains will no longer be favoured and new strains that accumulate so many mutations in the spike area as not to be recognised by the antibodies can have a chance. These strains with so many mutations in the spike area, most likely result less infective. This can give us a chance to control SARS-CoV-2, but evolution never stops. If we lower our guard prematurely, evolution can have an opportunity to produce new mutations with increased infectivity on resistant strains.

### 3. SARS-CoV-2 meta-population genetic structure and its consequences

Two opposed theoretical models have been proposed for the population’s genetic structure. In the classical model^84,85^ a population would have very little genetic variant. A more effective genotype would be the dominant. Most of the arising mutants would be deleterious and would go extinct by natural selection. At some point, a more effective new mutant would emerge and favoured by natural selection, would become dominant in the population.

In the second model, the balanced model^85^, there would be plenty genetic variant This variability would be maintained partly by natural selection, that in certain circumstances would favour one genotypes and in other circumstances to others by mechanisms like frequency dependent selection or environmental heterogeneity.

Elucidating if different organisms’ populations fit to one or the other model led to one of the most fruitful population genetics controversies^66,79,86^. It also helped to give birth to the neutral theory of molecular evolution^87–89^ that postulates that, although there is great genetic variant at a molecular level, natural selection is not capable of distinguishing much of it, consequently it fluctuates within the populations by mutation-drift.

It is not only a mere academic question; the genetic structure of a population also determines how the evolution of that population will be. Specifically, it predicts how the emergence and fixation of new mutant strains will be, and how genetic variant is maintained through time and space.

Massive sequencing in the SARS-CoV-2 meta-population shows the presence of an enormous genetic variant, with almost 300,000 different genetic variants, that quickly changes in space and time.

However, when unveiling the genetic structure of SARS-CoV-2’s population, it is paramount to consider that most of the mutants found are variants with no detectable biological effect whatsoever. In the eyes of natural selection, they are neutral. Although with our molecular tools can be detected, natural selection does not favour nor disfavour them. Only are maintained within the populations at a low frequency while others go extinct, luck determines their fate. If they happen in a super-spreader, that also infects other super-spreaders, the likelihood to continue in the population increases. In any case, most of them are lost after few generations and have no major consequences over COVID-19’s infectivity or lethality. They are, however, very useful for tracking contacts in medical practice.

In addition, deleterious genetic variants also arise from these neutral mutants. As they are unlikely to be detected, most of them disappear very quickly from the populations and are underestimated on the databases.

Mutants that apparently increase significantly viral infectivity also arise. For example, on https://www.gisaid.org^13^ we can follow the history of mutants as S477N, N439K or the N501Y variant, which seem to have some selective advantage and thank to that have significantly increased its frequency in the population. D614G strain, for example, emerged at the beginning of the COVID-19 pandemic in Europe, increased its frequency and was responsible of many infections during the first wave. A222V strain, that arise during the 2020 summer spread through Europe and increased its frequency during the second wave. At the end of 2020, United Kingdom reported a new variant VUI 202012/01 defined by multiple spike protein mutations (i.e., deletion 69-70, deletion 145, N501Y, A570D, D614G, P681H, T716I, S982A, D1118H), it become dominant in south east England and started spreading around the globe, In November the novel variant 501Y.V2, with triple spike receptor binding substitutions, was detected in South Africa and seems to be also increasing its frequency and spreading worldwide.

To have a better understanding of the SARS-CoV-2 population’s genetic structure, we can estimate the fraction of mutants with selective advantage respective to the neutral ones with selective disadvantage that really happen in the meta-population. Using the mutant surviving likelihood through time depending on its selective advantage or disadvantage (Eq. 1 to 7) and doing a sampling of the different predominant genetic variants in the UK and Europe shown on the databases through December 2020 to determine the fraction genetic variants favoured by natural selection respective to total genetic variants, is easy to estimate that for each of those that may have a selective advantage there are more than a thousand neutral or deleterious. In this way, SARS-CoV-2 meta-population structure resembles that proposed in the neutral evolution theory by Kimura (1968, 1979, 1989, 1991)^87–90^ and Ohta (1996)^91^.

We must not make the mistake of interpreting the population structure of SARS-CoV-2 under our technology perspective and not from the evolution perspective. In spite of that great variability at a molecular level, at an evolution level SARS-CoV-2 meta-population behaves as a population that follows the classical model. The vast majority of the mutations that arise become extinct sooner or later without reaching a sufficient frequency. Only a few, very infective, are favoured by selection and become dominant in the population for a length of time, until a new strain emerges that excels existing ones and natural selection favours to the top.

### 4. Muller’s ratchet, “mutational meltdown” and fundamental principle of natural selection

Nowadays Muller’s ratchet and mutational meltdown are ideas with ample prestige. A lot of theoretical and experimental work has been developed in the attempt of proving them^23,24,26,92–94^. In a meta-population of an RNA virus, as SARS-CoV-2 is, deleterious mutations should accumulate and with time should diminish the virus efficiency (Muller’s rachet) or even led it to extinction (mutational meltdown). It was indeed suggested to increase SARS-CoV-2 mutation rates to fight COVID-19 pandemic^28–30^.

But reality shows quite the opposite, SARS-CoV-2 is increasingly contagious because new strains emerge, originated by mutation, that are more effective and infectious than the ancestral ones they derive from.

We propose that the monumental size of SARS-CoV-2 populations, assures the occurrence of certain mutations with great selective advantage in coronavirus still with no deleterious mutations in its genome. Even being a very low likelihood event, the cosmic size of SARS-CoV-2 population makes it a sure event. New strains, originated by mutation, more effective and infectious than ancestral ones can still happen.

In any case and although not very fashionable, several theoretical models exist that can explain how microorganisms with no recombination could persist without suffering “mutational meltdown”^95–99^. There are also laboratory experiments where the opposite to expected in Muller’s rachet can be observed. Using clone cultures of microorganisms that reproduce asexually without recombination. Some aliquots of the ancestral cultures of several strains of these microorganisms where long kept frozen while allowing normal reproduction of the other aliquots. After many generations, when Muller’s rachet should have worked, the frozen cultures where thawed. It was verified that cultures that were allowed to reproduce normally had more fitness than ancestral ones, the opposite to expected according to Muller’s rachet^100^. Anyway, the existence in nature of millions of species that reproduce asexually, without recombination, is the best proof that Muller’s rachet can be avoided.

In the same sense, SARS-CoV-2 evolution seems to adjust better to fundamental principle of natural selection proposed by Fisher (1930)^37^. Fisher tried to explain evolution with his theorem. According to this principle, natural selection makes populations maximize their biological effectiveness through time. On his firsts formulations of his principle, he wrote that populations maximized their “intrinsic rate of natural increase”. Later he defined it using the more specific expression of the Malthusian fitness as: “the net reproductive rate of a population”. Nearly four decades later, Li (1967a, b)^101,102^ proved the fundamental principle of natural selection with a sophisticated mathematical reasoning.

Fundamental principle of natural selection predicts that the evolution of the SARS-CoV-2 population will make it every time more infective.

## Data Availability

All data used for the completion of this manuscript is of public access via the internet

## Competing Interest Statement

The authors have declared no competing interest.

## Notes

### Funding Statement

No funding was needed to complete this manuscript

### Author Declarations

No approval was needed for the completion of this work

